# Placenta-associated adverse pregnancy outcomes in women experiencing mild or severe hyperemesis gravidarum – a systematic review and meta-analysis

**DOI:** 10.1101/2022.11.09.22281870

**Authors:** Tilda Moberg, Lennart Van der Veeken, Emma Persad, Stefan R. Hansson, Matteo Bruschettini

## Abstract

**Background:** Nausea and vomiting in pregnancy (NVP) affects 50-80% of pregnant women and is correlated to the level of human chorionic gonadotropin (hCG). Hyperemesis gravidarum (HG) is a severe condition, with an incidence of 0.2-1.5%, characterized by consistent nausea, vomiting, weight loss and dehydration continuing after the second trimester.

**Aim:** The aim of this systematic review was to investigate a potential correlation between NVP or HG with adverse pregnancy outcomes and hCG levels.

**Method:** A systematic search in PubMed, Embase and CINAHL Complete was conducted. Studies on pregnant women with nausea in the first or second trimester, reporting either pregnancy outcomes or levels of hCG were included. The primary outcomes were preterm delivery (PTD), preeclampsia, miscarriage, and fetal growth restriction. Risk of bias was assessed using ROBINS-I. The overall certainty of evidence was assessed using GRADE.

**Results:** The search resulted in 2023 potentially relevant studies; 23 were included. The evidence was uncertain for all outcomes, however women with HG had a tendency to have an increased risk for preeclampsia [odds ratio (OR) 1.18, 95% confidence of interval (CI) 1.03 to 1.35], PTD [OR 1.35, 95% CI 1.13 to 1.61], small for gestational age (SGA) [OR 1.24, 95% CI 1.13 to 1.35], and low birth weight (LBW) [OR 1.35, 95% CI 1.26 to 1.44]. Further, a higher foetal female/male ratio was observed [OR 1.36, 95% CI 1.15 to 1.60]. Meta-analyses were not performed for women with NVP; however, most of these studies indicated that women with NVP have a lower risk for PTD and LBW and a higher risk for SGA, and a higher fetal female/male ratio.

**Conclusion:** There may be an increased risk in women with HG and a decreased risk in women with NVP for adverse placenta-associated pregnancy outcomes, however the evidence is very uncertain.

PROSPERO: CRD42021281218

## Introduction

Nausea and vomiting (NVP) in pregnancy affects about 50-80% of women. Usually, the nausea begins in the 4^th^ gestational week (gw) and resolves before the 20^th^ gw (1, 2). The levels of hCG increase rapidly during the first weeks of pregnancy, typically peaking around 9 gw, consistent with the peak of nausea and vomiting. The levels of hCG and the occurrence of nausea and vomiting are higher in molar- and multiple pregnancies, indicating that hCG correlates with these symptoms (3). The levels of hCG have also been shown to be higher in hyperemesis gravidarum (HG) (4).

Hyperemesis gravidarum is a severe condition characterized by consistent vomiting and nausea, weight loss, dehydration, ketonuria and electrolyte imbalances, with an incidence of 0.2-1.5%. The cause of HG remains unknown (5-7). It is classified as either mild or severe. Severe cases often require hospital care with intravenous fluids, antiemetics and sometimes parental nutrition. In fact, it is one of the common reasons for a pregnant woman to be admitted for hospital care (3, 8). Research studies have shown that HG has an increased risk for placenta-associated complications, such as fetal growth restriction (FGR), preterm delivery (PTD), preeclampsia, placentae abruptio and small for gestational age (SGA), when compared to less severe nausea (3, 9-11). Furthermore, it has been shown that there is an increased ratio between female and male foetuses in women admitted to the hospital due to HG (5, 8, 12, 13). The mechanism for the correlation between fetal sex and HG is unclear, but it could be related to higher levels of hCG and different placental functions deriving from sexual dimorphism (12).

Women with NVP have been shown to have a 50-75% reduced risk for miscarriage, supporting the hypothesis that nausea is a sign of a well-functioning placenta (14, 15). A dysfunctional placenta can result in a number of severe conditions including preeclampsia, intrauterine fetal death (IUFD), miscarriage, PTD, FGR and placental abruptio (3, 9). Preeclampsia affects 3-7% of pregnant women and causes 18% of maternal deaths worldwide, particularly in developing countries (16).

Many women suffer from nausea in pregnancy and there does not seem to be a consensus on any correlation between pregnancy induced nausea and adverse pregnancy outcomes. The aim of this systematic review was therefore to investigate if NVP or HG correlates with adverse pregnancy outcomes and if the levels of hCG differ between these groups.

## Methods

The protocol of this systematic review was registered with the international prospective register of systematic reviews, PROSPERO (CRD42021281218), prior to screening studies for inclusion (17). The protocol included relevant information including the review question, search strategy, eligibility criteria, primary and secondary outcomes, how to assess risk of bias, how to perform quality assessment, which data to extract and strategy for data synthesis.

### Eligibility criteria

We included studies of pregnant women with NVP or HG in the first or second trimester; we excluded studies of women with pre-existing systemic diseases such as liver-, kidney- or thyroid disease, diabetes, and hyperemesis in the third trimester. We included randomised controlled trials, non-randomised controlled trials, case-control studies, and cohorts. We excluded conference abstracts, reviews, case reports and other study designs. A cohort study was defined as an observational study with one group only, in this review women with either NVP or HG. A case control study was defined as a study comparing two groups, in this review women with NVP or HG compared to women without these conditions. The primary pregnancy outcomes in this review were miscarriage (fetal death before gw 20), preeclampsia, FGR (fetal estimated weight below the 10th percentile with a retardation in growth) and PTD (delivery before gw 37). The secondary outcomes were SGA (weight below the 10th percentile), LBW (birth weight below 2500 g), IUFD (intrauterine death after gw 28), placentae abruptio, fetal sex, rate of multiple pregnancies, congenital malformations, and admission to neonatal intensive care unit (NICU). Additionally, one included outcome not stated in the protocol was the level of hCG in the maternal blood. Studies that reported neither hCG levels nor any type of pregnancy outcome were excluded.

### Search

A search strategy was developed in collaboration with an information specialist at the Lund University library to identify all studies on pregnant women with nausea, vomiting or HG, reporting pregnancy outcomes or levels of hCG in blood were included. The databases PubMed, Embase and CINAHL Complete were searched for eligible studies by using free text and Medical Subject Headings, described in S1 Appendix. No restrictions regarding language, publication year or publication status were applied. The search was conducted on 28 September 2021.

### Study selection and data extraction

The studies were screened by two independent researchers using Covidence, an online tool for screening and data extraction in systematic reviews (18). Conflicts were solved by a third party or by reaching consensus between the researchers. Translators were contacted to translate studies in languages other than English, if necessary. Title, author, year, country, study design, number of participants, trimester and outcomes were extracted and reported in a table. Odds Ratio (OR) were extracted for each outcome and reported in a table. If a study did not report OR, it was calculated using Review Manager 5.4 (RevMan) from the number of events and the total number of participants (19).

### Risk of bias

Risk of bias was assessed for each study by two independent researchers using Risk Of Bias In Non-Randomized Studies - of Interventions (ROBINS-I) (20). The eight domains assessed for risk of bias were: bias due to; I) confounding, II) selection of participants, III) classification of interventions, IV) deviation from intended interventions, V) missing data, VI) measurement of outcomes, VII) selection of the reported results and VIII) overall risk of bias. The risk could be low, moderate, serious, or critical. The overall risk of bias was set at the same level as the domain with the highest risk. The confounding factors considered were pre-gestational diseases (diabetes, hypertension, liver-, kidney-, thyroid- and autoimmune disease), preeclampsia, eclampsia, HELLP (Hemolysis, Elevated Liver enzymes and Low Platelets) syndrome, gestational trophoblastic disease, multiple pregnancy, and molar pregnancy.

### Overall certainty of the evidence

The guidance for “Grading of Recommendations Assessment, Development and Evaluation” (GRADE) was used to evaluate the overall certainty of evidence for the four primary outcomes: preeclampsia, PTD, miscarriage and FGR, and for the two secondary outcomes reported by most studies: SGA and LBW (21). Women with NVP and women with HG were assessed together. Each outcome was assessed for risk of bias, inconsistency, indirectness, imprecision, and publication bias.

### Data analysis

Data was entered into RevMan, and OR with 95% confidence intervals (CI) were calculated for the dichotomous outcomes. Meta-analyses were performed for the outcomes with at least three studies, and forest plots were generated. For the outcomes reported in fewer than three studies, the study results were compared to each other and described in a narrative text. Women with NVP and with HG were analysed in different meta-analyses for the individual outcomes. Mild and severe HG were analysed together. Sensitivity analysis was performed for the studies with moderate or serious/critical risk of bias. Subgroups were created for each outcome, dividing the studies into women with NVP and women with HG. Heterogeneity was measured by I^2^ and calculated in RevMan.

## Results

### Search results

The search resulted in 2023 studies; 109 duplicates were removed. The title and abstract screening resulted in 325 potentially relevant studies. A *post hoc* decision was made to only assess studies describing pregnant women with NVP or HG that reported either hCG levels or pregnancy outcomes. This resulted in 56 studies that were screened in full text by two authors. Seven of them had no full text available and were identified as “awaiting classification”. Out of the 26 excluded studies, 15 did not report relevant outcomes, five were reviews, four did not study pregnant women with nausea, two were duplicates. This resulted in a total of 23 included studies (3-5, 8-11, 15, 22-36) (Fig 1).

**Fig 1.**
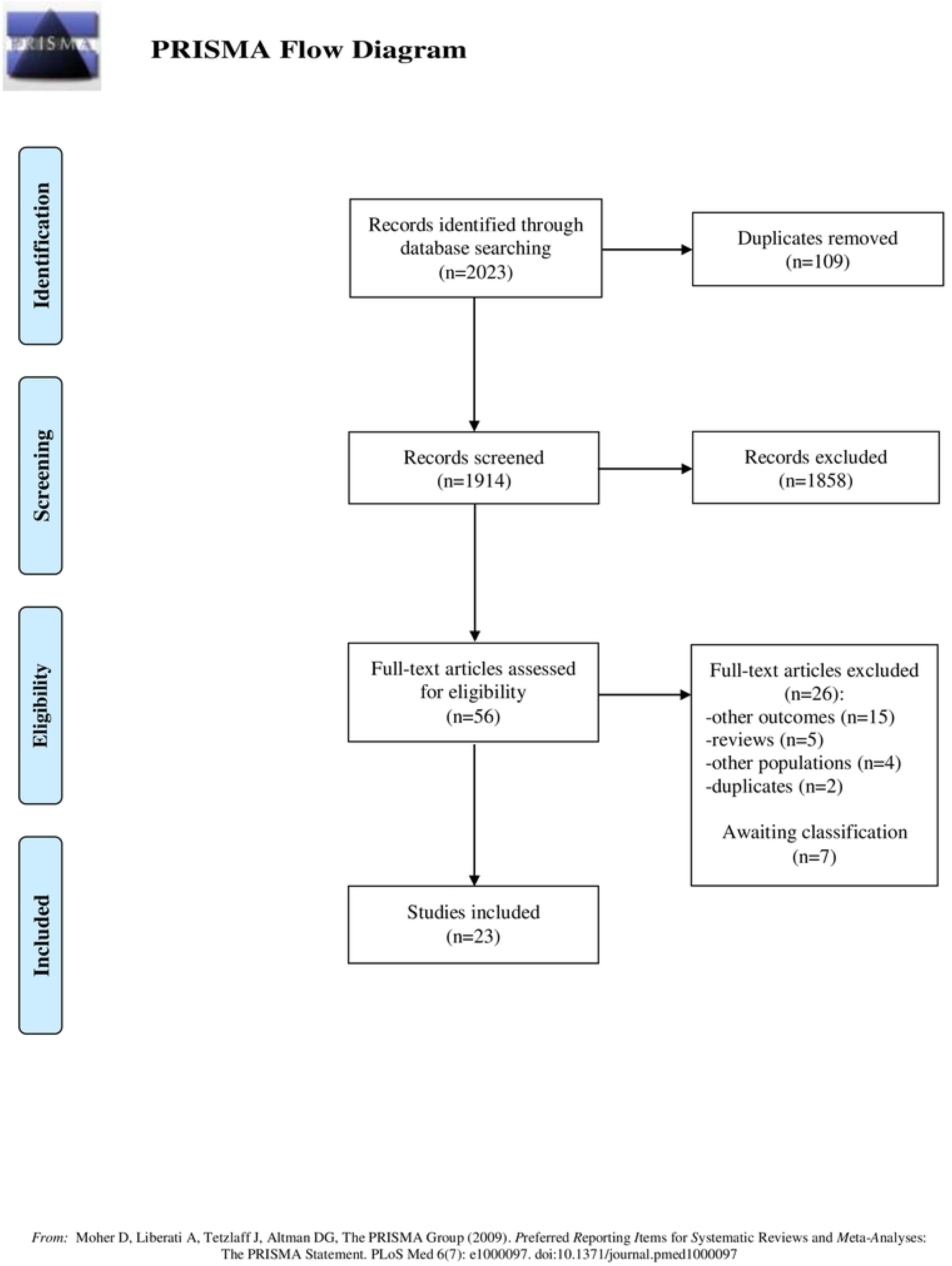
Flow chart. Flow chart of the study selection process.

### Included studies

Of the 23 included studies, 20 were case control studies and three were cohort studies. Of the included studies, 12 were prospective and 11 were retrospective. Fourteen studies included women with HG, two studies included women with nausea, two studies included women with vomiting, two studies included women with nausea and vomiting, two studies included two variables: one with nausea and one with nausea and vomiting, and one study reported the use of antiemetics (Table 1). One study was translated from Persian, and one was translated from French.

**Table 1.** Table of characteristics.

| Title | Study id | Year of enrolment | Country | Ethnicity | Participants | Trimester | Pregnancy outcome / hCG |
| --- | --- | --- | --- | --- | --- | --- | --- |
| Hormone profile of Kuwaiti women with hyperemesis gravidarum | Al-Yatama 2002 | 2000 | Kuwait | Kuwaitis, Arabs, Asians, Other | 50 women with vomiting and 50 women without vomiting | 1st & 2nd | hCG |
| Use of antiemetic drugs during pregnancy in Sweden | Asker 2005 | 1995-2002 | Sweden | Unclear | 29,804 women using antiemetics, 635 768 not using antiemetics | 1st, 2nd & 3rd | PTD, SGA, LBW, congenital malformations, fetal sex, rate of multiple pregnancies |
| Hyperemesis gravidarum and risks of placental dysfunction disorders: a population-based cohort study | Bolin 2013 | 1997-2009 | Sweden | Not reported | 1,156,050 women with HG | 1st & 2nd | Preeclampsia, PTD, SGA, IUFD, ablatio placentae |
| Pregnancy complications and birth outcomes among women experiencing nausea only or nausea and vomiting during pregnancy in the Norwegian Mother and Child Cohort Study | Chortatos 2015 | 1999-208 | Norway | Not reported | 20,371 with nausea, 17,070 with nausea and vomiting and 14,234 without nausea or vomiting | 1st & 2nd | Preeclampsia, PTD, SGA, LBW, congenital malformations, fetal sex |
| Association between severe nausea and vomiting in pregnancy and lower rate of preterm births | Czeizel 2004 | 1980-1996 | Hungary | European | 3,869 women with nausea and vomiting and 34,282 women without nausea and vomiting | 1st & 2nd | PTD, LBW, fetal sex |
| Hormonal and psychological factors in nausea and vomiting during pregnancy | Dekkers 2020 | Not reported | Netherlands | Not reported | 1,682 women with nausea | 1st | hCG |
| Outcomes of Pregnancies Complicated by Hyperemesis Gravidarum | Dodds 2006 | 1988-2002 | Canada | Not reported | 1,270 women with HG and 154,821 women without HG | 1st & 2nd | PTD, SGA, LBW |
| Antihistamines and other prognostic factors for adverse outcome in hyperemesis gravidarum | Fejzo 2013 | 2007-2011 | USA | Caucasian | 254 women with HG and 308 women without HG | 1st | PTD, SGA, congenital malformations, fetal sex |
| Adverse Maternal and Birth Outcomes in Women Admitted to Hospital for Hyperemesis Gravidarum: a Population-Based Cohort Study | Fiaschi 2018 | 1997-2012 | England | White, Black and white, Asian, Black, Chinese, Other | 118,197 women with HG and 8,093,653 women without HG | 1st & 2nd | Preeclampsia, PTD, SGA, LBW, IUFD, ablatio placentae, NICU admission |
| Pregnancy nausea related to women's obstetric and personal histories | Gadsby 1997 | Not reported | UK | Unclear | 363 women with nausea | Unclear | Fetal sex |
| Increased concentration of the free beta-subunit of human chorionic gonadotropin in hyperemesis gravidarum | Goodwin 1994 | Not reported | USA | Not reported | 39 women with HG and 23 women without HG | 1st & 2nd | hCG |
| The role of chorionic gonadotropin in transient hyperthyroidism of hyperemesis gravidarum | Goodwin 1992 | 1990 | USA | Not reported | 57 women with HG and 57 women without HG | 1st | hCG |
| [Hyperemesis gravidarum and pregnancy outcomes] | Hastoy 2015 | 2006-2009 | France | Not reported | 197 women with HG and 392 women without HG | 1st & 2nd | FGR, PTD, SGA, LBW, fetal sex |
| Hyperemesis gravidarum: is an ultrasound scan necessary? | Kirk 2006 | 2001-2006 | UK | Not reported | 286 women with HG and 286 women without HG | 1st | Miscarriage, rate of multiple pregnancies |
| Hyperemesis gravidarum and placental dysfunction disorders | Koudijs 2016 | 2012-2014 | Indonesia | Not reported | 400 women with HG and 1,833 women without HG | 1st & 2nd | Miscarriage, preeclampsia, SGA, LBW, IUFD |
| Outcomes of pregnancies complicated by hyperemesis gravidarum | Kuru 2012 | 2003-2011 | Turkey | Not reported | 72 women with HG and 89 women without HG | 1st & 2nd | PTD, SGA, IUFD, fetal sex |
| Immunologic and biochemical factors in hyperemesis gravidarum with or without hyperthyroxinemia | Leylek 1999 | Not reported | Turkey | Not reported | 30 women with HG and 15 women without HG | 1st | hCG |
| Maternal characteristics largely explain poor pregnancy outcome after hyperemesis gravidarum | Roseboom 2011 | 2000-2006 | Netherlands | Non-Western and Western | 5,207 women with nausea and vomiting and 76,279 women without nausea and vomiting | Unclear | PTD, SGA, IUFD, fetal sex, NICU admission |
| Pregnancy outcome in hyperemesis gravidarum and the effect of laboratory clinical indicators of hyperemesis severity | Tan 2007 | 2004-2005 | Kuala Lumpur | Malay, Chinese, Indian, Others | 166 women with HG and 498 women without HG | 1st & 2nd | Miscarriage, PTD, LBW, IUFD |
| Adverse pregnancy outcomes in women with nausea and vomiting of pregnancy | Temming 2014 | 2004-2011 | USA | African-American, American | 5,207 women with nausea and vomiting and 76,279 women without nausea and vomiting | Not reported | PTD, SGA, LBW, IUFD, fetal sex |
| Is the nausea and vomiting of early pregnancy really fetoprotective? | Weigel 2006 | Not reported | Ecuador | Unclear | 129 women with nausea, 373 women with nausea and vomiting and 131 women without nausea and vomiting | 1st & 2nd | Miscarriage, PTD, SGA, LBW, congenital malformations |
| Evaluating complications of pregnancy in patients with hyperemesis gravidarum | Yazdani 2010 | Not reported | Iran | Not reported | 70 women with HG and 100 women without HG | 1st & 2nd | Preeclampsia, PTD |
| Severe vomiting during pregnancy: antenatal correlates and fetal outcomes | Zhang 1991 | 1986-1987 | China | Not reported | 201 women with vomiting and 1,666 women without vomiting | Not reported | Preeclampsia, PTD, SGA, LBW, fetal sex |

Characteristics of the included studies. HG=hyperemesis gravidarum, PTD=preterm delivery, SGA=small for gestational age, LBW=low birth weight, IUFD=intrauterine fetal death, GD=gestational diabetes, PIH=pregnancy induced hypertension, GW=gestational week, NICU=neonatal intensive care unit, FGR=fetal growth restriction, LGA=large for gestational age.

### Risk of bias of the included studies

Most of the studies (13 studies) were considered to have an overall moderate risk of bias (Fig 2). Seven studies were considered to have an overall serious risk of bias and three studies were considered to have an overall critical risk of bias. None of the studies were considered to have an overall low risk of bias. All the studies were considered to have at least moderate risk of bias due to confounding factors for nausea. All the studies had a low risk of bias in the selection of participants and in the measurements of outcomes.

**Fig 2.**
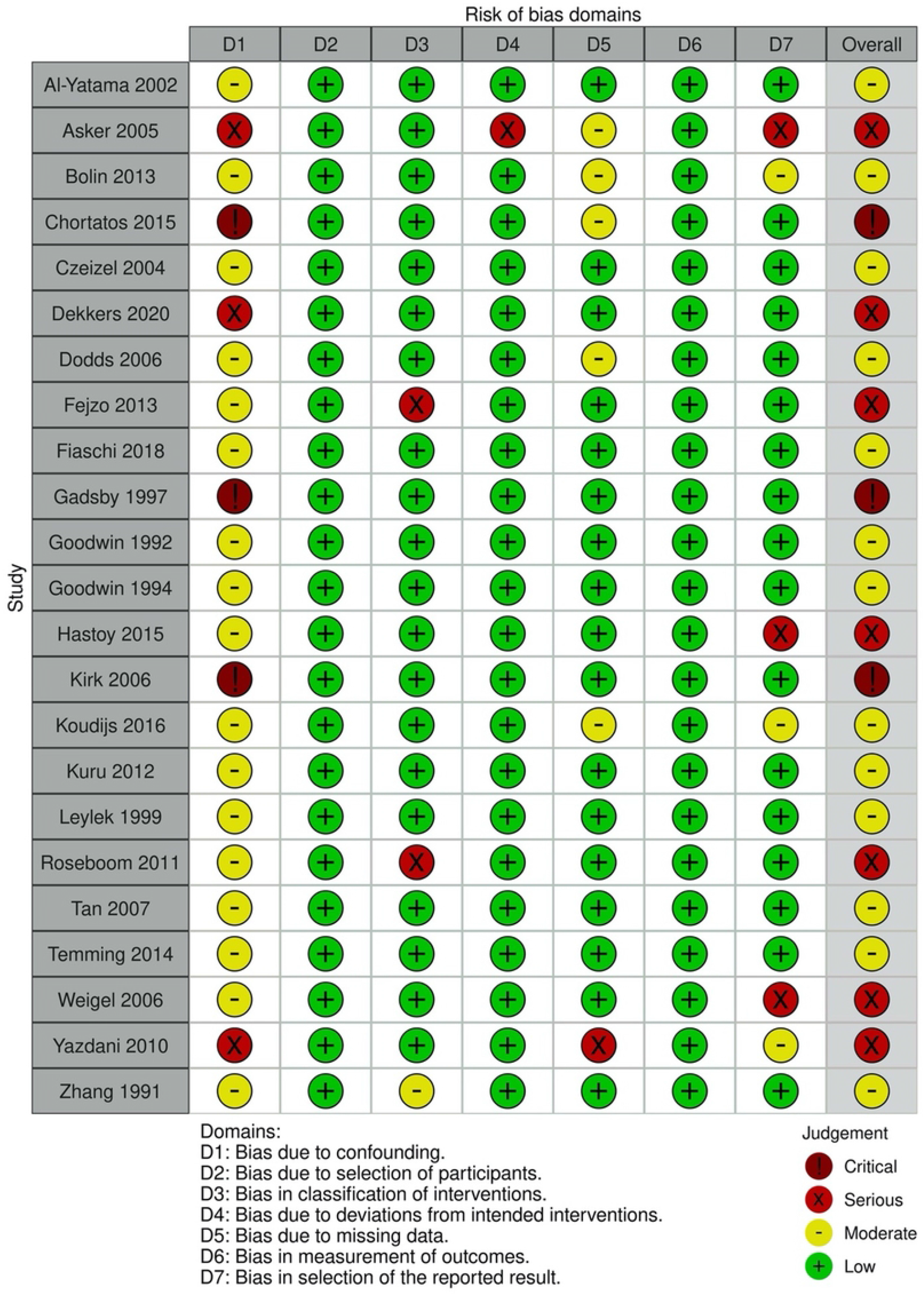
Risk of bias assessment. Risk of bias assessment for all included studies. The risk of bias for each domain and the overall risk of bias are shown. The overall risk of bias was set at the same level as the domain with the highest risk of bias.

### Data analysis

Odds ratios for each study and outcome are reported in Table 2. Most outcomes were not reported by a sufficient number of studies to be pooled in a meta-analysis and were therefore analysed individually. Sensitivity analysis did not reduce the heterogeneity remarkably and is not shown in the final analysis. The study by Weigel *et al*. (34), reporting PTD, miscarriage, SGA, low birth weight and congenital malformations, could not be included in the meta-analyses since the numbers for the control group was not reported.

**Table 2.**
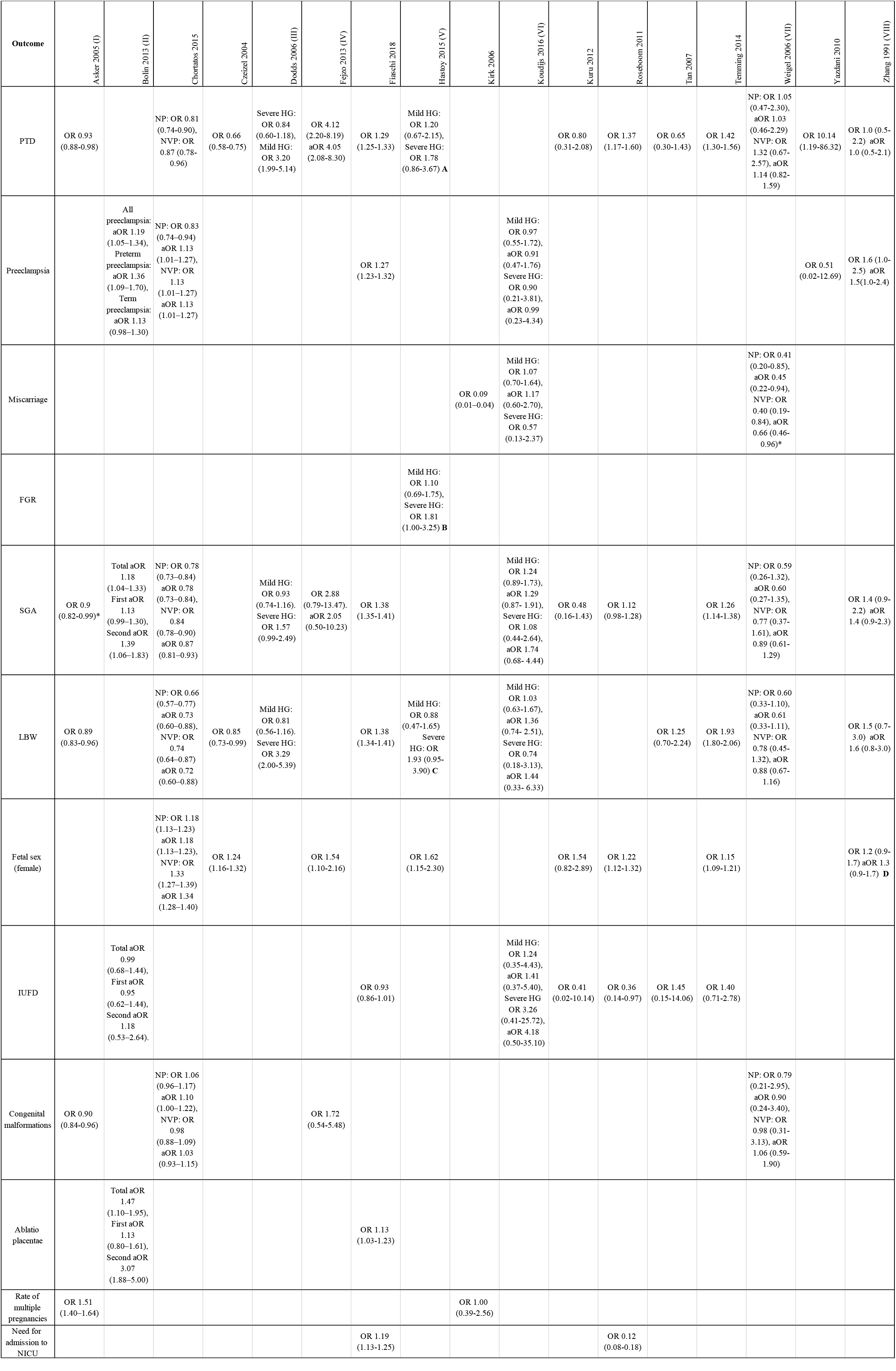
Table of outcomes.

Odds ratio with 95% confidence intervals, OR (CI), are shown for all the outcomes. PTD=preterm delivery, FGR=fetal growth restriction, SGA=small for gestational age, LBW=low birth weight, IUFD=intrauterine fetal death, OR=odds ratio. aOR=adjusted OR. CI=confidence interval, HG=hyperemesis gravidarum, NP=nausea during pregnancy, NVP=nausea and vomiting during pregnancy, gw=gestational week, BMI=body mass index. (I) Adjustments were made for maternal age, parity, BMI, height, smoking, cohabitation with infant’s father, infant’s sex, mother’s country of birth and years of formal education, presence of hyperthyreosis, pregestational diabetes, chronic hypertension, and year of birth of infant. (II) Preeclampsia: Adjusted for age, BMI, smoking, parity, education, and gender. SGA: Adjusted for age, BMI, smoking, education, and gender. LBW: Adjusted for age, BMI, smoking, parity, education, gender, gestational length, and energy intake. Fetal sex: Adjusted for age, BMI, smoking, parity, and education. (III) Mild HG: Weight gain ≥7kg. Severe HG: Weight gain <7kg. (IV) Unclear what is adjusted for. (V) A: Adjusted for maternal age, parity and smoking. B: Adjusted for smoking, body mass index and maternal blood pressure. C: Adjusted for smoking and maternal body mass index. Mild HG: weight gain ≥7 kg. Severe HG: weight gain < 7kg. (VI) Adjusted model: adjusted for socio-economic status (as reflected by income), smoking status, gravidity, maternal age and pre-pregnancy BMI. Mild HG: weight loss ≥5% of pre-pregnancy weight. Severe HG: weight loss <5% of pre-pregnancy weight. (VII) Adjusted for maternal age, neighborhood altitude, periconceptual use of prenatal tobacco, alcohol, antiemetic drugs, and vitamin-mineral supplements. SGA and low birth weight are in addition adjusted for gestational age. (VIII) Adjusted for maternal age, chronic illnesses, and paternal smoking. D=Adjusted for gravidity primae, maternal age >30 years, chronic hypertension, chronic liver disease, congenital or rheumatic heart diseases, chronic renal illness, paternal smoking.

Preterm delivery (PTD) was reported in 14 studies with a total of 9,054,950 participants, of whom Fiaschi *et al*. (11), contributed 6,835,060. The results were pooled in two different meta-analyses, one for NVP and one for HG. The meta-analysis for HG resulted in OR 1.35, 95% CI 1.13 to 1.61, indicating that women with HG have a higher risk for PTD (Fig 3). Due to the high heterogeneity, the studies on women with NVP could not be pooled in a meta-analysis. Three of these five studies showed a significantly lower risk for PTD in women with NVP, although the certainty of evidence is very low. One study showed a significant higher risk for PTD in women with NVP, and one study showed no difference. The study by Weigel *et al*. (34) was excluded due to missing data.

**Fig 3.**
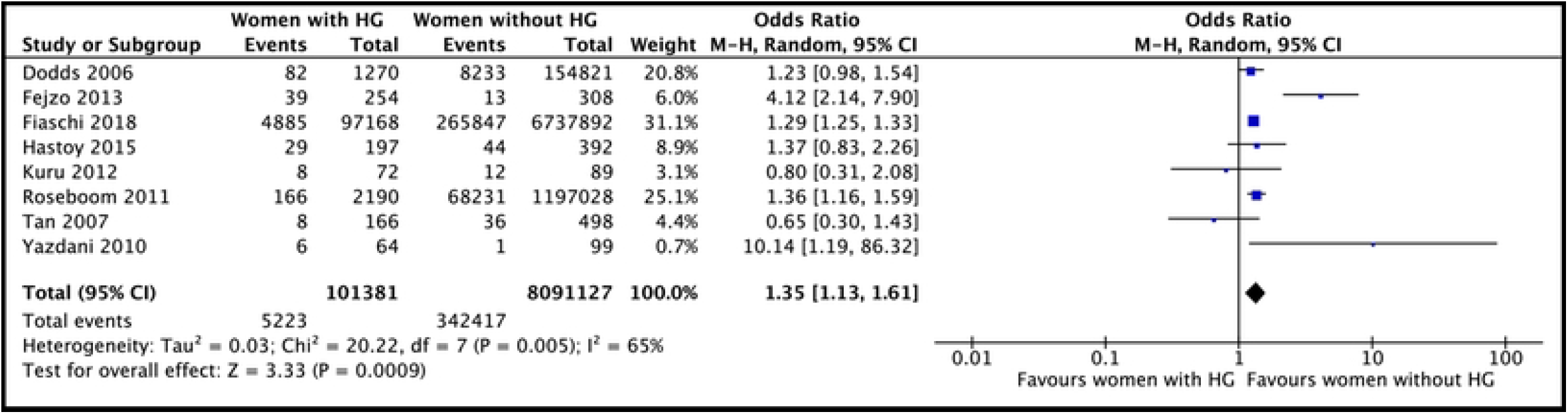
Meta-analysis for preterm delivery. Forest plot showing the meta-analysis for preterm delivery in women with HG. Odds ratios with 95% confidence intervals (CI) are reported. The heterogeneity is showed as the value of I2.

Preeclampsia was reported in six studies with a total of 9,421,054 participants. Four of these studies reported women with HG and the meta-analysis resulted in OR 1.18, 95% CI 1.03 to 1.35, suggesting that HG is a risk factor for preeclampsia (Fig 4). Since only two studies reported on women with NVP, a meta-analysis was not performed. One of the studies showed a significant higher risk for preeclampsia in women with NVP. The other study showed no difference.

**Fig 4.**
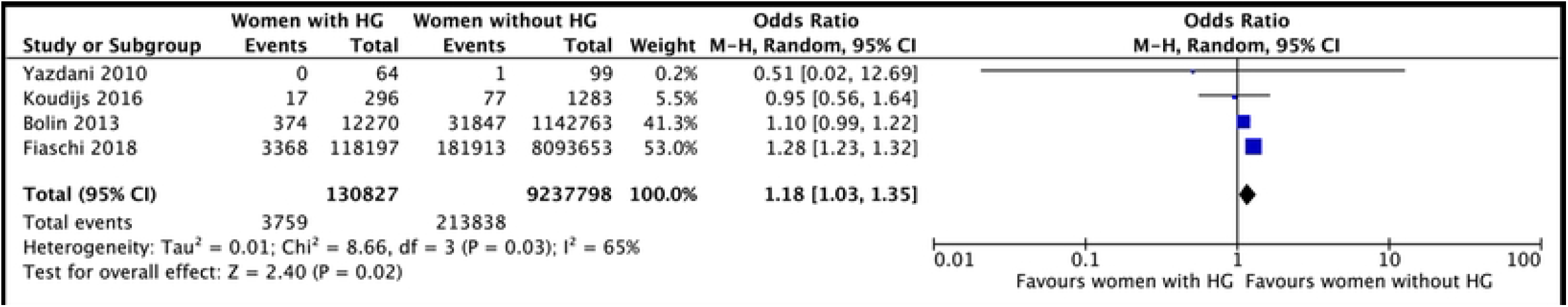
Meta-analysis for preeclampsia. Forest plot showing the meta-analysis for preeclampsia in women with HG. Odds ratios with 95% confidence intervals (CI) are reported. The heterogeneity is showed as the value of I^2^.

Miscarriage was reported in three studies including 3,438 women. A meta-analysis was not performed due to high heterogeneity and missing data. The study by Weigel *et al*. (34) did not report the numbers for the control group, hence no OR, anticipated absolute effect or event rates could be calculated. One study showed a significant decreased risk for miscarriage in women with HG, and the other study reported no difference. The study by Tan *et al*. (32) reported two cases of miscarriage but excluded them from their analysis.

Fetal growth restriction (FGR) was only reported in one study, Hastoy *et al*. (8). Its definition was translated to SGA in the English abstract but was then described as FGR in the rest of the French article. The total number of participants in this study was 589 and the results indicated an increased risk for FGR in women with HG.

Small for gestational age (SGA) was reported in 12 studies with a total number of 7,797,344 participants. Seven studies reported SGA in women with HG and were pooled in a meta-analysis, which resulted in OR 1.24, 95% CI 1.13 to 1.35, (Fig 5). Due to high heterogeneity, the studies on women with NVP could not be pooled in a meta-analysis. Three of four studies showed an increased risk for SGA in women with NVP, though only two significant. The fourth study showed a significant decreased risk for SGA in women with NVP.

**Fig 5.**
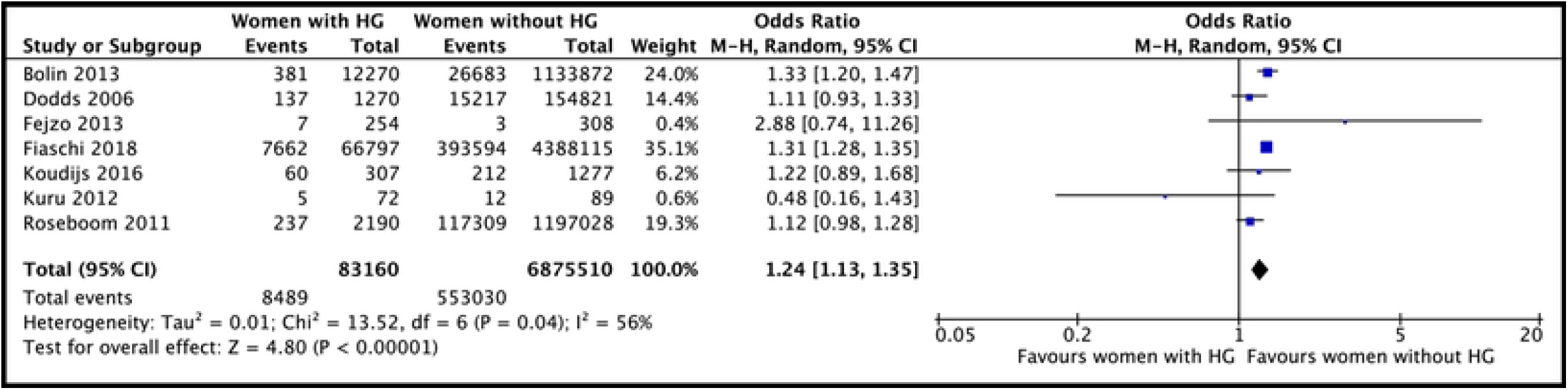
Meta-analysis for small for gestational age (SGA). Forest plot showing the meta-analysis for SGA in women with HG. Odds ratios with 95% confidence intervals (CI) are reported. The heterogeneity is showed as the value of I^2^.

Low birth weight (LBW) was reported in 11 studies with a total number of 7,824,780 participants. The studies reporting LBW in women with HG were pooled in a meta-analysis, which resulted in OR 1.35, 95% CI 1.26 to 1.44, (Fig 6). Due to high heterogeneity, the studies on women with NVP could not be pooled in a meta-analysis. Three of these five studies showed a significant reduced risk for LBW in women with NVP. The other two studies showed an increased risk for LBW in women without NVP, although only one was statistically significant. The study by Weigel *et al*. (34) was excluded due to missing data.

**Fig 6.**
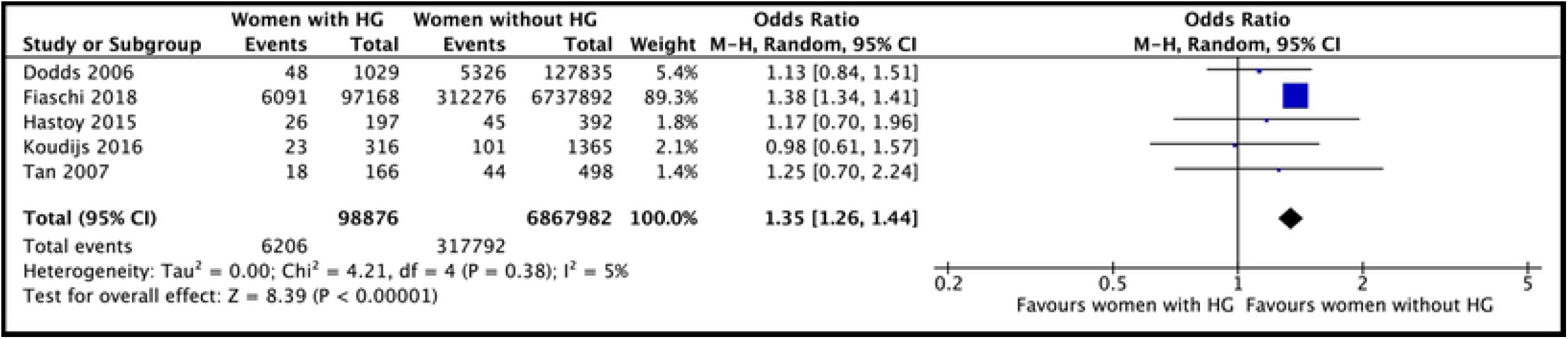
Meta-analysis for low birth weight (LBW). Forest plot showing the meta-analysis for LBW in women with HG. Odds ratios with 95% confidence intervals (CI) are reported. The heterogeneity is showed as the value of I^2^.

Fetal sex was reported in eight studies with a total number of 1,446,151 participants. The studies reporting women with HG were pooled in a meta-analysis, which resulted in OR 1.36, 95% CI 1.15 to 1.60, (Fig 7). Due to high heterogeneity, the studies on women with NVP could not be pooled in a meta-analysis. Three of four studies showed higher probability for the foetus being female in women with NVP. The other study showed a significant lower probability of the foetus being female in women with NVP, contributing to over half of the total number of participants.

**Fig 7.**
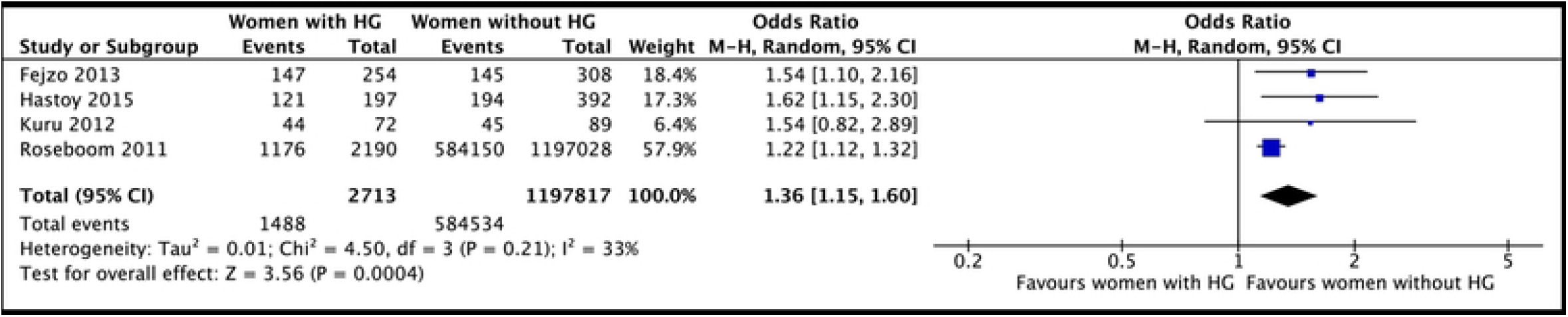
Meta-analysis for foetal sex (female). Forest plot showing the meta-analysis for foetal sex (female) in women with HG. Odds ratios with 95% confidence intervals (CI) are reported. The heterogeneity is showed as the value of I^2^.

Intrauterine fetal death (IUFD) was reported in seven studies with a total number of 10,650,249 participants. A meta-analysis was performed for the six studies reporting IUFD in women with HG, which resulted in OR 0.94, 95% CI 0.86 to 1.02, (Fig 8). The study by Fiaschi *et al*. (11) reported this outcome in two different groups: one group with only fetal deaths and one group where one foetus died, and one lived (multiple pregnancy). In the meta-analysis the groups were analysed together. There was only one study reporting IUFD in women with NVP, indicating a higher risk for IUFD for women with NVP, but the results were not significant.

**Fig 8.**
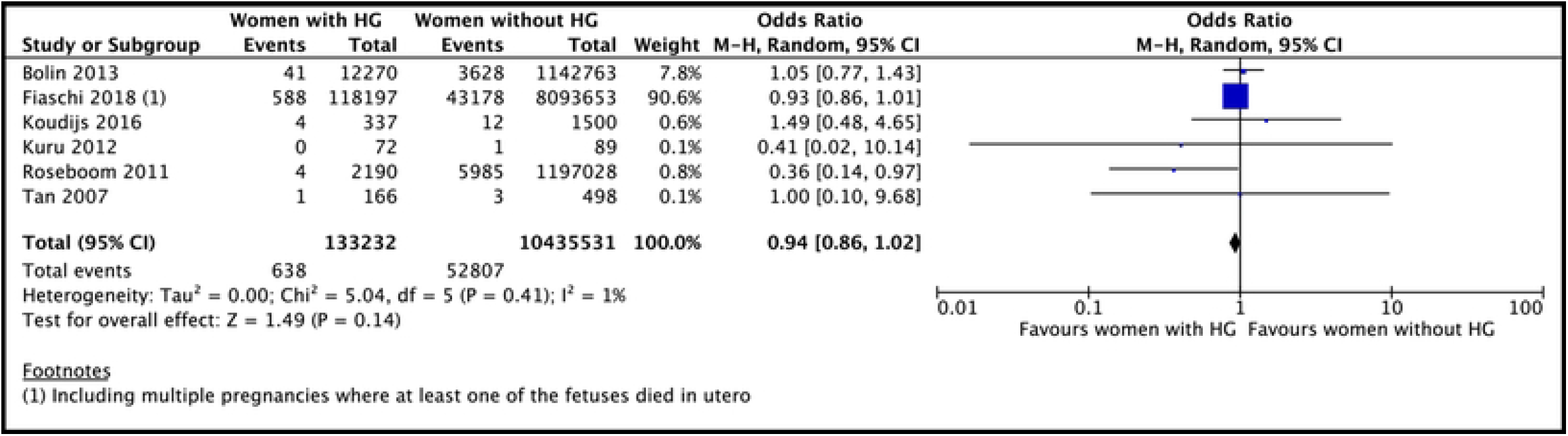
Meta-analysis for intrauterine foetal death (IUFD). Forest plot showing the meta-analysis for IUFD in women with HG. Odds ratios with 95% confidence intervals (CI) are reported. The heterogeneity is showed as the value of I^2^.

Congenital malformations were reported in four studies with a total of 704,208 participants. A meta-analysis was not performed due to high heterogeneity. Two of the studies showed a decreased risk for congenital malformations in women with NVP and two studies showed an increased risk for women with NVP or HG.

Placentae abruptio was reported in two studies with a total of 9,367,900 participants. A meta-analysis was not performed since the outcome was only reported in two studies. Both studies showed significant results of an increased risk for placentae abruptio in women with HG.

Rate of multiple pregnancies was reported by two studies with a total of 666,144 participants. A meta-analysis was not performed since the outcome was only reported in two studies. Both studies showed higher rates of multiple pregnancies in women with NVP or HG.

Need for admission to NICU was reported in two studies with a total of 9,411,068 participants. A meta-analysis was not performed since the outcome was only reported in two studies. Both studies showed higher rates of admission to NICU in women with HG.

### Human choriogonadotropin (hCG) levels

The five studies that reported levels of hCG in women with NVP or HG are described in Table 3. Four of them were case control studies and one was a cohort study. Since the cohort study did not have a control group, only the levels of hCG in hyperaemic patients are reported in Table 3. The studies measured hCG with different methods, at different gestational ages and reported the results in different units. Free ß-hCG was reported in three studies, intact hCG in three studies, and *α*-hCG in one study.

**Table 3.**
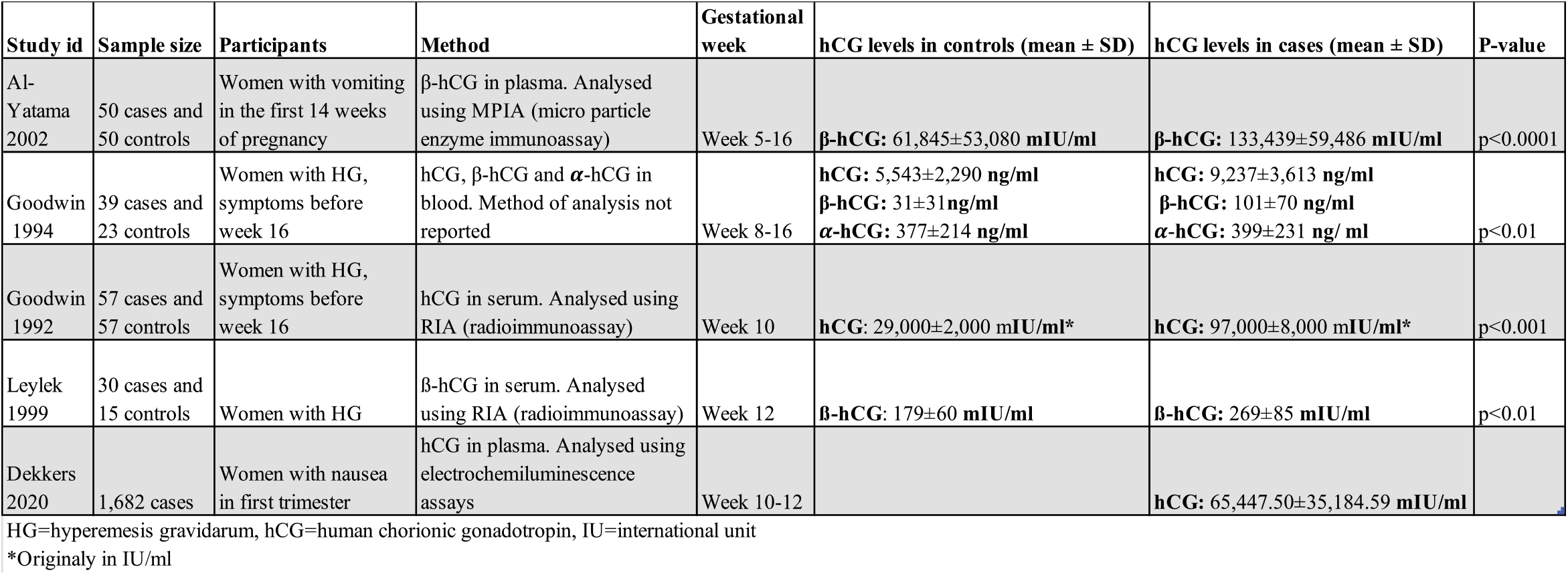
Levels of hCG.

Comparison of hCG levels, reported in five studies.

### GRADE assessment

Certainty of evidence for the four primary outcomes and for SGA and LBW was very low (see Table 4). All outcomes were downgraded for imprecision of the estimates. Miscarriage and FGR were downgraded also for serious risk of bias. Preeclampsia, PTD, SGA and LBW were downgraded for inconsistency. Study event rates and anticipated absolute effect are reported in the summary of findings table (Table 4).

**Table 4.** Summary of findings.

| Outcomes | Number of participants (studies) | Overall certainty of evidence (GRADE) | Relative effect OR (95% CI) | Anticipated absolute effects |  |
| --- | --- | --- | --- | --- | --- |
|  |  |  |  | Risk without nausea | Risk with nausea |
| Miscarriage | 3438 (3) | Very low ⊕○○○ (a) |  |  |  |
| Preeclampsia | 9421054 (6) | Very low ⊕○○○ (b) | 1.14 (0.97 to 1.33) | 23 per 1000 | 8 more per 1000 |
| Preterm delivery | 9054950 (14) | Very low ⊕○○○ (b) | 1.13 (0.94 to 1.36) | 43 per 1000 | 8 more per 1000 |
| Fetal growth restriction | 589 (1) | Very low ⊕○○○ (c) | 1.30 (0.87 to 1.93) | 217 per 1000 | 47 more per 1000 |
| Small for gestational age | 7796711 (12) | Very low ⊕○○○ (b) | 1.28 (0.95 to 1.73) | 75 per 1000 | 9 more per 1000 |
| Low birth weight | 7824780 (11) | Very low ⊕○○○ (b) | 1.04 (0.83 to 1.30) | 45 per 1000 | 3 more per 1000 |

(a) due to serious risk of bias, high heterogeneity and wide CI
(b) due to high heterogeneity and wide CI
(c) due to serious risk of bias and wide CI

Summary of findings and certainty of evidence assessment (GRADE). The four primary outcomes and two of the secondary outcomes (SGA and LBW) were assessed. Both women with NVP and with HG are included.

## Discussion

The aim of this systematic review was to explore if there are any correlations between NVP or HG and adverse placenta-associated outcomes. We found that there might be an association between HG and an increased risk for PTD, preeclampsia, SGA, and LBW, however the certainty of evidence was very low. Most studies indicated that women with NVP have a lower risk for PTD and LBW, but a higher risk for SGA. The fetal female/male ratio was in general higher in both women with NVP and HG.

Limitations of this review include the *post hoc* decision to not include the studies on hCG in correlation to adverse pregnancy outcomes, as specified in the review protocol (17). By including studies in all languages, not applying restrictions for publishing year and by performing an extensive search in three different large databases, the risk of missing potentially relevant studies was minimised. To reduce the risk of confounders, studies on women with intercurrent diseases were excluded. Several intercurrent diseases have been associated with an increased risk for HG, e.g., pre-gestational diabetes and thyroid diseases (5, 37). The confounding factors existing for nausea are difficult to adjust for in observational studies. Therefore, none of the studies were individually considered to have an overall low risk of bias due to confounding factors. One of the cohort studies reported the use of antiemetics in pregnant women, i.e., indirectly reporting on the symptom nausea. The study included women with prescriptions for antiemetics as cases and all other women as controls. Women buying antiemetics over the counter were not registered as cases, generating a higher risk of bias due to misclassifications of the interventions. Some of the studies did not report a clear definition for HG, also increasing the risk of bias for classification of interventions. ROBINS-I was used since it is the most suitable tool for assessing risk of bias in non-randomised studies. For many of the outcomes, the heterogeneity was high due to different study characteristics and diverse results. The imprecision was also high due to few studies reporting the outcome, a small number of participants in the included studies and broad CIs. The certainty of evidence according to GRADE was therefore considered very low for all the assessed outcomes. The uncertain evidence entails the greatest limitation of this review.

Other systematic reviews by Veenendaal *et al*. (38) and Varela *et al*. (39) studying women with HG and adverse pregnancy outcomes including PTD, LBW and SGA, have shown results in line ours, namely, an increased risk for adverse pregnancy outcomes in women with HG. The increased risk for these outcomes in women with HG could be a consequence of the systemic and metabolic effects generated by HG, negatively affecting the woman and the foetus. When studying mild and severe HG separately, an increased risk for adverse pregnancy outcome is particularly associated with the severe form of HG (8, 10, 40). Severe HG is associated with low caloric intake, weight loss, electrolyte imbalance and disturbances in glucose metabolism, reducing the availability of nutrients for the foetus as well. It is a state comparable to famine, where studies have shown that a reduction of intrauterine nutrition correlates to lower birth weight (41). Only one review, Koren *et al*. (42), was found that reported adverse pregnancy outcomes in women with NVP. They showed a reduced risk for miscarriage, congenital malformations and PTD; results that are in line with this review.

This systematic review also intended to investigate if there are correlations between levels of hCG and NVP or HG. The studies reporting levels of hCG were heterogeneous and reported different types of hCG (alfa-, beta- and intact hCG), with different analysing methods, at different gestational ages and in different units. As there is a lack of consensus regarding equivalent units, a comparable unit could not be generated. However, this was not a major limitation in the aspect of this review, since the comparison between cases and controls was the most important, showing higher levels of hCG in the cases. On the other hand, only five studies were found and most of them were published over 20 years ago, indicating that there is limited amount of ongoing research in this area. Although the levels of hCG in pregnant women have not changed throughout the years, more studies are needed to be able to draw well-grounded conclusions about to which extent the levels differ in women with HG.

It could be argued that high levels of hCG originate from a well-functioning placenta producing adequate levels and thereby ensuring a decreased risk for adverse pregnancy outcomes. Nausea has shown to be protective against miscarriage, and the levels of hCG seem to be higher in women experiencing nausea in early pregnancy (4, 14, 15). On the other hand, results from other studies have shown the opposite effect, indicating that too high levels of hCG are associated with adverse pregnancy outcomes. The levels of hCG have been shown to be higher in women with HG, in which the incidence of adverse pregnancy outcomes also is high, confirming the results in this review (43). In addition, hCG in combination with other biomarkers, such as pregnancy-associated plasma protein A (PAPP-A), alpha fetoprotein (AFP), inhibin-A and unconjugated estriol (uE3) is used in prenatal screening tests for chromosome abnormality, where high levels of hCG are indicative for, such as Down syndrome. However, there seems to be an important difference between milder nausea and HG. As shown in this review, the rate of adverse pregnancy outcomes seems to be reduced in women with NVP but increased in women with HG. Women with nausea might have a light increase of hCG that could indicate a well-functioning placenta. To date, this has seemingly not been studied specifically in women with NVP.

No other systematic review was found that reported the correlations for both NVP and HG, levels of hCG and pregnancy outcomes. The results of this review are important to help our understanding of the risks for adverse pregnancy outcomes in women with NVP or HG, which could contribute to a decreased morbidity and mortality for both the mother and the child. This review also highlights the need for further research in this area. It is important that future research further decreases the risk of confounders to make the results more reliable. For instance, by reporting data in women with intercurrent diseases separately. Future studies should also focus on studying HG stratified by mild and severe HG, given the results of a higher incidence of adverse pregnancy outcomes in women with severe HG.

## Conclusion

The evidence of this research, although uncertain, suggests that women with HG may have an increased risk for adverse placenta-associated outcomes whilst women with milder nausea may have a reduced risk. Both women with NVP and HG have a high female/male fetal ratio, suggesting an interesting sexual dimorphism. The levels of hCG were found to be higher in women with HG. Further research is needed to draw grounded conclusions about any correlations. Above all, studies with lower risk of bias are needed to improve the certainty of evidence.

## Data Availability

All relevant data are within the manuscript and its Supporting Information files.

## Acknowledgements

We thank Maria Björklund (Library and ICT services, Lund University, Sweden) for contributing to the search strategy and Ehsan Hedayati (Ahvaz, Iran) for translating a study from Persian into English.

## Notes

### Competing Interest Statement

The authors have declared no competing interest.

### Funding Statement

The authors received no specific funding for this work.

## References

1. Niebyl JR. Nausea and Vomiting in Pregnancy. New England Journal of Medicine. 2010;363(16):1544–50.

2. Jarnfelt-Samsioe A. Nausea and vomiting in pregnancy: a review. Obstet Gynecol Surv. 1987;42(7):422–7.

3. Koudijs HM, Savitri AI, Browne JL, Amelia D, Baharuddin M, Grobbee DE, et al. Hyperemesis gravidarum and placental dysfunction disorders. BMC Pregnancy Childbirth. 2016;16(1):374.

4. Goodwin TM, Hershman JM, Cole L. Increased concentration of the free beta-subunit of human chorionic gonadotropin in hyperemesis gravidarum. Acta Obstet Gynecol Scand. 1994;73(10):770–2.

5. Roseboom TJ, Ravelli AC, van der Post JA, Painter RC. Maternal characteristics largely explain poor pregnancy outcome after hyperemesis gravidarum. Eur J Obstet Gynecol Reprod Biol. 2011;156(1):56–9.

6. Tsang IS, Katz VL, Wells SD. Maternal and fetal outcomes in hyperemesis gravidarum. Int J Gynaecol Obstet. 1996;55(3):231–5.

7. Organization WH. ICD-10 : international statistical classification of diseases and related health problems : tenth revision, 2nd ed. : World Health Organization; 2004 [Available from: https://apps.who.int/iris/handle/10665/42980.

8. Hastoy A, Lien Tran P, Lakestani O, Barau G, Gérardin P, Boukerrou M. [Hyperemesis gravidarum and pregnancy outcomes]. J Gynecol Obstet Biol Reprod (Paris). 2015;44(2):154–63.

9. Bolin M, Akerud H, Cnattingius S, Stephansson O, Wikström A. Hyperemesis gravidarum and risks of placental dysfunction disorders: a population-based cohort study. BJOG: An International Journal of Obstetrics & Gynaecology. 2013;120(5):541–7.

10. Dodds L, Fell DB, Joseph KS, Allen VM, Butler B. Outcomes of Pregnancies Complicated by Hyperemesis Gravidarum. Obstetrics & Gynecology. 2006;107(2 Part 1):285–92.

11. Fiaschi L, Nelson-Piercy C, Gibson J, Szatkowski L, Tata LJ. Adverse Maternal and Birth Outcomes in Women Admitted to Hospital for Hyperemesis Gravidarum: a Population-Based Cohort Study. Paediatr Perinat Epidemiol. 2018;32(1):40–51.

12. Schiff MA, Reed SD, Daling JR. The sex ratio of pregnancies complicated by hospitalisation for hyperemesis gravidarum. Bjog. 2004;111(1):27–30.

13. Basso O, Olsen J. Sex ratio and twinning in women with hyperemesis or pre-eclampsia. Epidemiology. 2001;12(6):747–9.

14. Hinkle SN, Mumford SL, Grantz KL, Silver RM, Mitchell EM, Sjaarda LA, et al. Association of Nausea and Vomiting During Pregnancy With Pregnancy Loss: A Secondary Analysis of a Randomized Clinical Trial. JAMA Intern Med. 2016;176(11):1621–7.

15. Kirk E, Papageorghiou AT, Condous G, Bottomley C, Bourne T. Hyperemesis gravidarum: is an ultrasound scan necessary? Hum Reprod. 2006;21(9):2440–2.

16. Hansson SR, Nääv Å, Erlandsson L. Oxidative stress in preeclampsia and the role of free fetal hemoglobin. Frontiers in Physiology. 2015;5(516).

17. Moberg T HS, Van der Veeken L, Bruschettini M.. Correlation between pregnancy induced nausea and placenta associated outcomes. PROSPERO CRD420212812182021 [Available from: Available from: https://www.crd.york.ac.uk/prospero/display_record.php?ID=CRD42021281218.

18. Covidence systematic review software VHI, Melbourne, Australia. Available at https://www.covidence.org.

19. Collaboration TC. Review Manager (RevMan) Version 5.4. 2020.

20. Sterne JA, Hernán MA, Reeves BC, Savović J, Berkman ND, Viswanathan M, et al. ROBINS-I: a tool for assessing risk of bias in non-randomised studies of interventions. Bmj. 2016;355:i4919.

21. GRADEpro GDT [Computer program]. Version accessed 16 March 2018. Hamilton (ON): McMaster University (developed by Evidence Prime) Aago. [

22. Al-Yatama M, Diejomaoh M, Nandakumaran M, Monem RA, Omu AE, Al Kandari F. Hormone profile of Kuwaiti women with hyperemesis gravidarum. Arch Gynecol Obstet. 2002;266(4):218–22.

23. Asker C, Norstedt Wikner B, Källén B. Use of antiemetic drugs during pregnancy in Sweden. Eur J Clin Pharmacol. 2005;61(12):899–906.

24. Chortatos A, Haugen M, Iversen PO, Vikanes Eberhard-Gran M, Bjelland EK, et al. Pregnancy complications and birth outcomes among women experiencing nausea only or nausea and vomiting during pregnancy in the Norwegian Mother and Child Cohort Study. BMC Pregnancy and Childbirth. 2015;15(1).

25. Czeizel AE, Puhó E. Association between severe nausea and vomiting in pregnancy and lower rate of preterm births. Paediatr Perinat Epidemiol. 2004;18(4):253–9.

26. Dekkers GWF, Broeren MAC, Truijens SEM, Kop WJ, Pop VJM. Hormonal and psychological factors in nausea and vomiting during pregnancy. Psychol Med. 2020;50(2):229–36.

27. Fejzo MS, Magtira A, Schoenberg FP, MacGibbon K, Mullin P, Romero R, et al. Antihistamines and other prognostic factors for adverse outcome in hyperemesis gravidarum. Eur J Obstet Gynecol Reprod Biol. 2013;170(1):71–6.

28. Gadsby R, Barnie-Adshead AM, Jagger C. Pregnancy nausea related to women’s obstetric and personal histories. Gynecol Obstet Invest. 1997;43(2):108–11.

29. Goodwin TM, Montoro M, Mestman JH, Pekary AE, Hershman JM. The role of chorionic gonadotropin in transient hyperthyroidism of hyperemesis gravidarum. J Clin Endocrinol Metab. 1992;75(5):1333–7.

30. Kuru O, Sen S, Akbayır O, Goksedef BP, Ozsürmeli M, Attar E, et al. Outcomes of pregnancies complicated by hyperemesis gravidarum. Arch Gynecol Obstet. 2012;285(6):1517–21.

31. Leylek OA, Toyaksi M, Erselcan T, Dokmetas S. Immunologic and biochemical factors in hyperemesis gravidarum with or without hyperthyroxinemia. Gynecol Obstet Invest. 1999;47(4):229–34.

32. Tan PC, Jacob R, Quek KF, Omar SZ. Pregnancy outcome in hyperemesis gravidarum and the effect of laboratory clinical indicators of hyperemesis severity. J Obstet Gynaecol Res. 2007;33(4):457–64.

33. Temming L, Franco A, Istwan N, Rhea D, Desch C, Stanziano G, et al. Adverse pregnancy outcomes in women with nausea and vomiting of pregnancy. J Matern Fetal Neonatal Med. 2014;27(1):84–8.

34. Weigel MM, Reyes M, Caiza ME, Tello N, Castro NP, Cespedes S, et al. Is the nausea and vomiting of early pregnancy really feto-protective? Journal of Perinatal Medicine. 2006;34(2):115–22.

35. Yazdani S, Bouzari Z, Faghani S. Evaluating complications of pregnancy in patients with hyperemesis gravidarum. Feyz Journal of Kashan University of Medical Sciences. 2010;14(4):426–30.

36. Zhang J, Cai WW. Severe vomiting during pregnancy: antenatal correlates and fetal outcomes. Epidemiology. 1991;2(6):454–7.

37. Fell DB, Dodds L, Joseph KS, Allen VM, Butler B. Risk Factors for Hyperemesis Gravidarum Requiring Hospital Admission During Pregnancy. Obstetrics & Gynecology. 2006;107(2 Part 1):277–84.

38. Veenendaal MV, van Abeelen AF, Painter RC, van der Post JA, Roseboom TJ. Consequences of hyperemesis gravidarum for offspring: a systematic review and meta-analysis. Bjog. 2011;118(11):1302–13.

39. Varela P, Deltsidou A. Hyperemesis gravidarum and neonatal outcomes: A systematic review of observational studies. Taiwanese Journal of Obstetrics and Gynecology. 2021;60(3):422–32.

40. Gross S, Librach C, Cecutti A. Maternal weight loss associated with hyperemesis gravidarum: a predictor of fetal outcome. Am J Obstet Gynecol. 1989;160(4):906–9.

41. Lumey LH. Reproductive outcomes in women prenatally exposed to undernutrition: a review of findings from the Dutch famine birth cohort. Proc Nutr Soc. 1998;57(1):129–35.

42. Koren G, Madjunkova S, Maltepe C. The protective effects of nausea and vomiting of pregnancy against adverse fetal outcome—A systematic review. Reproductive Toxicology. 2014;47:77–80.

43. Lepage N, Chitayat D, Kingdom J, Huang T. Association between second-trimester isolated high maternal serum maternal serum human chorionic gonadotropin levels and obstetric complications in singleton and twin pregnancies. Am J Obstet Gynecol. 2003;188(5):1354–9.

