## Supplementary material for "Placenta-associated adverse pregnancy outcomes in women experiencing mild or severe hyperemesis gravidarum – a systematic review and meta-analysis": S Appendix

### **S1 Appendix**

**Search strategy**

**PubMed**

**#1:** Search: (((((((pregnancy complication*[Title/Abstract])) OR (pregnancy outcome*[Title/Abstract])) OR (miscarriage[Title/Abstract])) OR (pregnancy loss[Title/Abstract])) OR (intrauterine death[Title/Abstract] OR intra-uterine death[Title/Abstract] OR fetal death[Title/Abstract] OR IUFD[Title/Abstract] OR intrauterine fetal death[Title/Abstract]) OR (stillbirth[Title/Abstract]) OR (ablatio[Title/Abstract] OR placental abruption[Title/Abstract] OR abruptio placentae[Title/Abstract] OR IUGR[Title/Abstract] OR intrauterine growth restriction[Title/Abstract] OR intra-uterine growth restriction[Title/Abstract] OR fetal growth retardation[Title/Abstract] OR fetal growth restriction[Title/Abstract] OR FGR[Title/Abstract] OR preterm delivery[Title/Abstract] OR pre-term delivery[Title/Abstract] OR prematurity[Title/Abstract] OR premature birth[Title/Abstract] OR neonatal morbidity[Title/Abstract] OR neonatal mortality[Title/Abstract] OR Fetal sex[Title/Abstract] OR sex of fetus[Title/Abstract] OR sex of foetus[Title/Abstract]) OR (mola[Title/Abstract]) OR (mola hydatidosa[Title/Abstract])) OR (recurrent miscarriage[Title/Abstract])) OR (((((((("pregnancy complications"[MeSH Terms]) OR ("abruptio placentae"[MeSH Terms])) OR ("abortion, spontaneous"[MeSH Terms])) OR (abortion, threatened[MeSH Terms])) OR (fetal growth retardation[MeSH Terms])) OR (infant, small for gestational age[MeSH Terms]) OR ("fetal death"[MeSH Terms]) OR (premature birth[MeSH Terms]) OR ("eclampsia"[MeSH Terms]) OR (hellp syndrome[MeSH Terms]) OR (recurrent miscarriage[MeSH Terms]) OR ("stillbirth"[MeSH Terms])) OR ("abortion, habitual"[MeSH Terms]))) Filters: Humans

**443 225 records**

**#2:** Search: (("placenta"[MeSH Terms]) OR ("pre eclampsia"[MeSH Terms]) OR (eclampsia[MeSH Terms])) OR (HELLP[Title/Abstract]) OR (Hemolysis Elevated Liver enzymes Low Platelet[Title/Abstract]) OR (placenta[Title/Abstract]) OR (placental function[Title/Abstract]) OR (placental disorder*[Title/Abstract]) OR (placental dysfunction[Title/Abstract]) OR (preeclampsia[Title/Abstract]) OR (pre-eclampsia[Title/Abstract]) OR (previous preeclampsia[Title/Abstract] OR hypert*[Title/Abstract]) OR ((placenta*[Title/Abstract] OR preeclampsia[Title/Abstract] OR pre-eclampsia) AND (diabetes[Title/Abstract] OR hypert*[Title/Abstract])) Filters: Humans

**583 019 records**

**#3:** Search: ((chorionic gonadotropin[MeSH Terms]) OR (hyperemesis gravidarum[MeSH Terms]) OR (hyperemesis[Title/Abstract]) OR (hyperemesis gravidarum[Title/Abstract])) OR ((Pregnanc* OR gestation*[Title/Abstract]) AND (vomit*[Title/Abstract] OR nausea[Title/Abstract])) OR (hCG[Title/Abstract] OR human chorionic gonadotropin[Title/Abstract] OR chorionic gonadotropin[Title/Abstract]) AND (humans[Filter]) Filters: Humans

**38 787 records**

**#4:** #1 AND #2 AND #3

**2 476 records**

**#5:** Search: (("review"[Publication Type]) OR ("case reports"[Publication Type])) OR ("letter"[Publication Type]) Filters: Humans

**4 971 960 records**

**#6:** #4 NOT #5

**1 623 records**

**CINAHL Complete**

**#1**: pregnancy complication* OR pregnancy outcome* OR miscarriage OR pregnancy loss OR intrauterine death OR intra-uterine death OR fetal death OR IUFD OR intrauterine fetal death OR stillbirth OR ablatio OR placental abruption OR abruptio placentae OR IUGR OR intrauterine growth restriction OR intra-uterine growth restriction OR fetal growth retardation OR fetal growth restriction OR FGR OR preterm delivery OR pre-term delivery OR prematurity OR premature birth OR neonatal morbidity OR neonatal mortality OR Fetal sex OR sex of fetus OR sex of foetus OR mola OR mola hydatidosa OR recurrent miscarriage OR pregnancy complication* OR pregnancy outcome* OR miscarriage OR pregnancy loss OR intrauterine death OR intra-uterine death OR fetal death OR IUFD OR intrauterine fetal death OR stillbirth OR ablatio OR placental abruption OR abruptio placentae OR IUGR OR intrauterine growth restriction OR intra-uterine growth restriction OR fetal growth retardation OR fetal growth restriction OR FGR OR preterm delivery OR pre-term delivery OR prematurity OR premature birth OR neonatal morbidity OR neonatal mortality OR Fetal sex OR sex of fetus OR sex of foetus OR mola OR mola hydatidosa OR recurrent miscarriage

**157 141 records**

**#2**: HELLP OR Hemolysis Elevated Liver enzymes Low Platelet OR placenta OR placental function OR placental disorder* OR placental dysfunction OR preeclampsia OR pre-eclampsia OR previous preeclampsia OR ((placenta* OR preeclampsia OR pre-eclampsia) AND (diabetes OR hypert*)) OR HELLP OR Hemolysis Elevated Liver enzymes Low Platelet OR placenta OR placental function OR placental disorder* OR placental dysfunction OR preeclampsia OR pre-eclampsia OR previous preeclampsia OR ((placenta* OR preeclampsia OR pre-eclampsia) AND (diabetes OR hypert*))

**27 074 records**

**#3**: hyperemesis OR hyperemesis gravidarum OR ((Pregnanc* OR gestation*) AND (vomit* OR nausea)) OR hCG OR human corionic gonadotropin OR chorionic gonadotropin OR hyperemesis OR hyperemesis gravidarum OR ((Pregnanc* OR gestation*) AND (vomit* OR nausea)) OR hCG OR human chorionic gonadotropin OR chorionic gonadotropin

**6 911 records**

**#4:** #1 AND #2 AND #3

**357 records**

**#5:** letters OR case report OR review

**859 844 records**

**#6:** #4 NOT #5

**268 records**

**#7:** #6 + Humans

**133 records**

**Embase**

**#1**: 'pregnancy complication'/exp OR 'solutio placentae'/exp OR 'abruptio placentae complications'/exp OR 'spontaneous abortion'/exp OR 'imminent abortion'/exp OR 'intrauterine growth retardation'/exp OR 'small for date infant'/exp OR 'fetus death'/exp OR 'prematurity'/exp OR 'hellp syndrome'/exp OR 'eclampsia'/exp OR 'recurrent miscarriage'/exp OR 'stillbirth'/exp OR 'recurrent abortion'/exp

**386 317 records**

**#2**: 'pregnancy complication*':ab,ti OR 'pregnancy outcome*':ab,ti OR miscarriage:ab,ti OR 'pregnancy loss':ab,ti OR 'intrauterine death':ab,ti OR 'intra-uterine death':ab,ti OR 'fetal death':ab,ti OR iufd:ab,ti OR 'intrauterine fetal death':ab,ti OR stillbirth:ab,ti OR ablatio:ab,ti OR 'placental abruption':ab,ti OR 'abruptio placentae':ab,ti OR iugr:ab,ti OR 'intrauterine growth restriction':ab,ti OR 'intra-uterine growth restriction':ab,ti OR 'fetal growth retardation':ab,ti OR 'fetal growth restriction':ab,ti OR fgr:ab,ti OR 'preterm delivery':ab,ti OR 'pre-term delivery':ab,ti OR prematurity:ab,ti OR 'premature birth':ab,ti OR 'neonatal morbidity':ab,ti OR 'neonatal mortality':ab,ti OR 'fetal sex':ab,ti OR 'sex of fetus':ab,ti OR 'sex of foetus':ab,ti OR mola:ab,ti OR 'mola hydatidosa':ab,ti OR 'recurrent miscarriage':ab,ti

**166 813 records**

**#3**: 'eclampsia'/exp OR 'preeclampsia'/exp OR 'placenta'/exp

**143 527 records**

**#4**: hellp:ab,ti OR 'hemolysis elevated liver enzymes low platelet':ab,ti OR placenta:ab,ti OR 'placental function':ab,ti OR 'placental disorder*':ab,ti OR 'placental dysfunction':ab,ti OR preeclampsia:ab,ti OR 'pre eclampsia':ab,ti OR 'previous preeclampsia':ab,ti OR ((placenta*:ab,ti OR preeclampsia:ab,ti OR 'pre eclampsia':ab,ti) AND (diabetes:ab,ti OR hypert*:ab,ti))

**127 013 records**

**#5:** 'chorionic gonadotropin'/exp OR 'hyperemesis gravidarum'/exp

**56 516 records**

**#6**: hyperemesis:ab,ti OR 'hyperemesis gravidarum':ab,ti OR ((pregnanc*:ab,ti OR gestation:ab,ti) AND (vomit:ab,ti OR nausea:ab,ti)) OR hcg:ab,ti OR 'chorionic gonadotropin':ab,ti OR 'human chorionic gonadotropin':ab,ti

**49 722 records**

**#7**: #1 OR #2

**450 965 records**

**#8**: #3 OR #4

**185 714 records**

**#9**: #5 OR #6

**75 372 records**

**#10**: #7 AND #8 AND #9

**2 218 records**

**#11**: #10 AND 'human'/de

**2 017 records**

**#12**: 'conference abstract':it OR ((review OR case) AND report)

**7 005 681 records**

**#13**: #10 AND 'human'/de NOT ('conference abstract':it OR ((review OR case) AND **report**))

**1 279 records**

**#14**: #13 AND [embase]/lim NOT ([embase]/lim AND [medline]/lim)

**269 records**
